# Timing of Illness Onset Influences Leukocyte Destruction in Sepsis

**DOI:** 10.1101/2025.06.28.25330461

**Authors:** Yoshitomo Morinaga, Motohiro Sekino, Kazuto Tsuruda, Hiroo Hasegawa, Tetsuya Hara, Katsunori Yanagihara

## Abstract

**Background:** The clinical significance of leukocyte destruction morphology in routine peripheral blood smears remains underexplored. This study aimed to systematically quantify leukocyte destruction and correlate these findings with clinical course, etiology, and outcome in patients with suspected sepsis.

**Methods:** This was a retrospective observational study involving 30 intensive care unit (ICU) patients with suspected sepsis. Peripheral blood smear findings were evaluated chronologically, with the day of blood culture collection designated as Day 0, followed by four distinct phases. Leukocyte destruction was defined as clear evidence of cytoplasmic or nuclear dissolution. Correlations were assessed with 28-day prognosis, etiology (infectious vs. non-infectious), and symptom onset (acute [no preceding symptoms before admission] vs. non-acute [preceding symptoms present]).

**Results:** Leukocyte destruction showed no significant difference concerning 28-day mortality or etiology. However, it was significantly more frequent in acute onset cases within the first 48 hours (7.0% vs. 4.1%, p<0.05). Conversely, cases with toxic granulation were significantly more common in non-acute onset cases within the first 48 hours (70% vs. 14%, p<0.01).

**Discussion:** Leukocyte destruction in acute onset and toxic granulation in non-acute onset offer potential diagnostic insights. These findings suggest uncharacterized leukocyte maturation or differentiation pathways influenced by illness timing, warranting further research for biomarker discovery and pathophysiology.

## Introduction

Morphological destruction of leukocytes, visible as cytoplasmic or nuclear disintegration in peripheral blood smears, is sometimes encountered; however, it is rarely quantified or systematically studied in clinical hematology. While such changes may reflect mechanical artifact, increasing evidence suggests that they are associated with underlying pathophysiological processes, especially in the context of severe infection or acute stress. Despite the routine use of blood smears in clinical practice, the diagnostic and prognostic value of leukocyte destruction morphology remains unclear.

Recent advances in neutrophil biology have revealed that neutrophil extracellular traps (NETs)—web-like structures composed of chromatin and antimicrobial proteins—play a critical role in host defense and immunopathology [1]. NETs formation can be triggered by both infectious and sterile inflammatory stimuli, and their excessive production has been linked to thrombosis [2], and organ dysfunction in sepsis [3, 4], COVID-19 [4, 5], and other acute illnesses [6]. However, most studies have relied on molecular or imaging approaches, and few have addressed whether NETs-like morphology or other forms of leukocyte destruction can be reliably detected and quantified in routine blood smears.

To date, there have been no original studies systematically quantifying leukocyte destruction morphology in the peripheral blood of patients with suspected sepsis and comparing these findings to other acute clinical presentations. The present study addresses this gap by providing a detailed morphological and temporal analysis of leukocyte destruction in critically ill patients, evaluating its association with clinical course, etiology, and outcome, and discussing its relationship to emerging concepts in neutrophil biology.

## Materials and Methods

### Patient Selection and Clinical Data

This retrospective observational study was conducted at Nagasaki University Hospital and included patients admitted to the intensive care unit (ICU) between November 2014 and May 2015. The study was approved by the ethics committee of Nagasaki University Hospital (approval number, 14102792).

Eligible patients were those who underwent blood culture testing due to suspected sepsis during their ICU stay. Written informed consent was obtained from all participants or their legal surrogates prior to inclusion. The final study cohort consisted of 30 patients, comprising 21 males and 9 females, with a mean age of 64.4 years (±16.0).

### Blood Sample Collection and Smear Preparation

Peripheral blood samples were collected from each patient during their ICU admission, using EDTA-2K as an anticoagulant to preserve cellular morphology. For each sample, a peripheral blood smear was prepared and stained with the May-Giemsa method, which enables detailed morphological evaluation of leukocytes. The day of blood culture collection was designated as Day 0 to standardize the temporal analysis across patients.

### Morphological Assessment and Definition of Leukocyte Destruction

The observation period was divided into four phases: Phase 1 (Day 0–2), Phase 2 (Day 3–5), Phase 3 (Day 6–8), and Phase 4 (Day 9–14). For each phase, blood smears were prepared and subjected to microscopic examination by experienced clinical laboratory scientists. However, not all patients had samples available for every designated phase, primarily due to the practical limitations of routine clinical workflow, such as weekend, staff availability, or patient discharge from the ICU.

In each smear, two independent counts of 100 leukocytes were performed, and the mean values for each cell type and morphological abnormality—including cytoplasmic and nuclear leukocyte destruction—were calculated (Fig 1A-E). In addition to leukocyte destruction, other morphological features such as vacuolization, toxic granulation, left shift, and Döhle bodies were systematically recorded (Fig 1G-I). Leukocyte destruction was defined as clear evidence of cytoplasmic or nuclear dissolution, distinct from simple apoptotic or necrotic changes.

**Figure 1.**
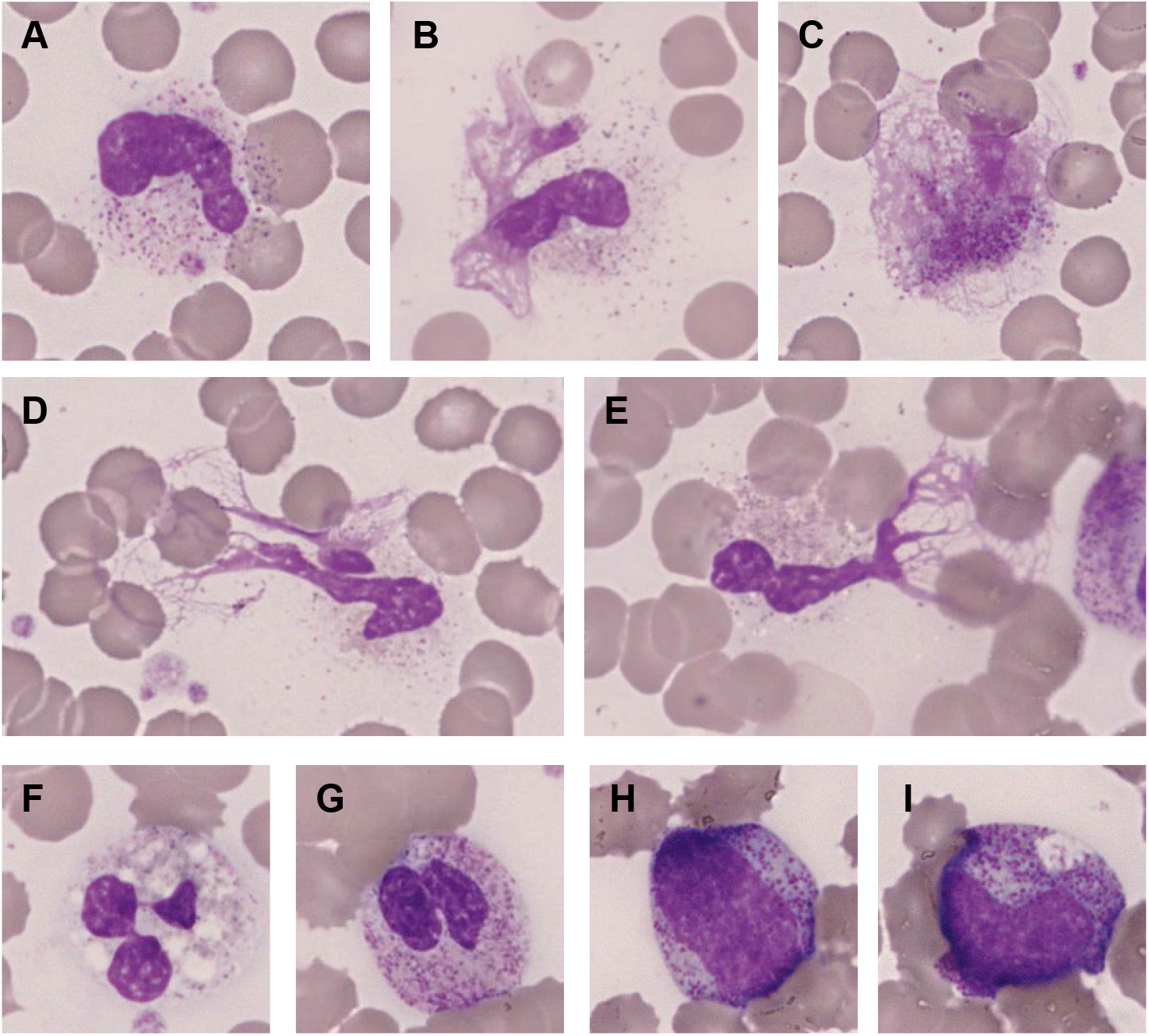
Representative images of leukocyte morphological features. **(A-E)** Images illustrating leukocyte destruction, characterized by clear evidence of cytoplasmic disintegration or nuclear fragmentation, as defined in this study. **(F-I)** Examples of other morphological abnormalities: (F) vacuolization, (G) toxic granulation, (H) left shift (immature neutrophil forms), and (I) Döhle bodies.

### Clinical Grouping

For clinical correlation, patients were categorized based on three criteria. First, prognosis was determined by patient outcome within 28 days of ICU admission, classifying them into a survivor group (survived at least 28 days) or a non-survivor group (died within 28 days). Second, cases were categorized as either infectious or non-infectious acute illness where no clear signs of infection were confirmed. The latter specifically included conditions such as severe pancreatitis, acute myocardial infarction, trauma, and burns. Patients were grouped as having acute onset if they presented with no preceding symptoms before ICU admission, or non-acute onset if they had experienced preceding symptoms prior to admission.

### Statistical Analysis

Statistical analysis was performed to evaluate the relationships between leukocyte destruction and the clinical variables described above. The Mann-Whitney U test was used for comparisons of continuous variables between two groups, and the chi-squared test was applied for categorical data. For time-series data, statistical analysis was conducted only for Phase 1, where a sufficient sample size was ensured. A p-value of less than 0.05 was considered statistically significant. All analyses were conducted using GraphPad Prism (version 10, GraphPad software, CA).

## Results

### Leukocyte Destruction and Clinical Associations

For clinical correlation, the mean proportion of leukocyte destruction in Phase 1 (Day 0-2) was analyzed in relation to various clinical variables.

For prognostic associations, no statistically significant difference in leukocyte destruction was found between survivors and non-survivors (p=0.56, Fig 2A). The mean proportion of leukocyte destruction in Phase 1 was 4.3 ± 0.8 % (n = 22, mean ± standard error of the mean) in survivors and 5.0 ± 1.3 % (n = 8) in non-survivors.

**Figure 2.**
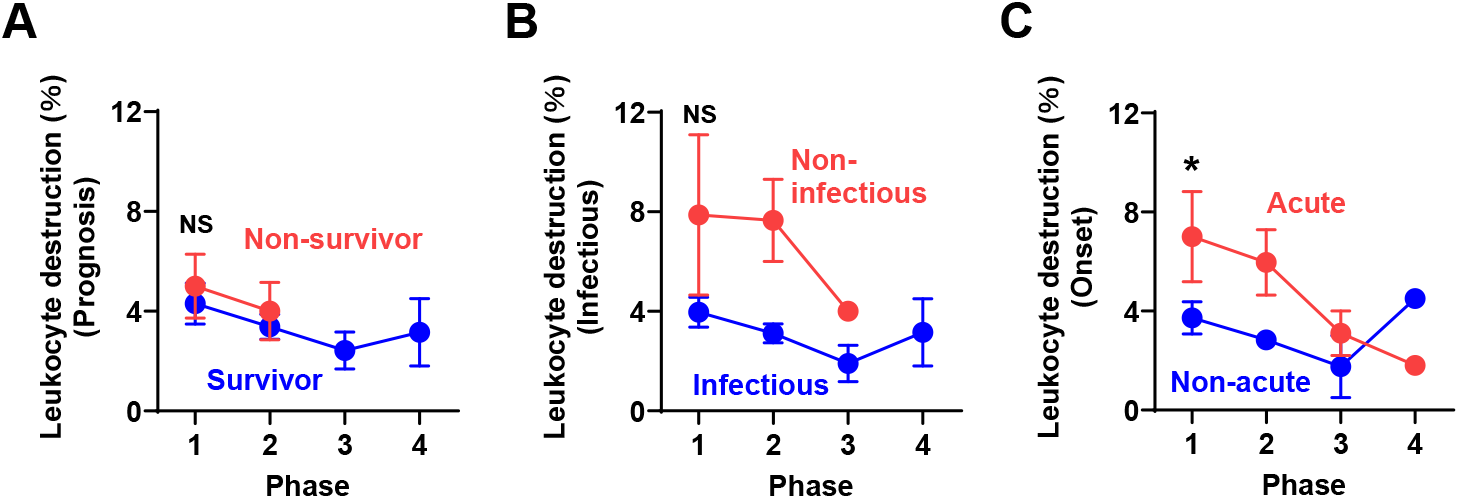
Leukocyte destruction over time stratified by clinical associations. The proportion of leukocyte destruction (%) is shown across four observation phases (Phase 1: Day 0–2; Phase 2: Day 3–5; Phase 3: Day 6–8; Phase 4: Day 9–14). Data are presented as mean ± standard error of the mean. **(A)** Prognostic association: Comparison of leukocyte destruction between 28-day survivors (blue) and non-survivors (red). **(B)** Etiological association: Comparison of leukocyte destruction between patients with infectious etiology (blue) and non-infectious acute illness (red). **(C)** Onset association: Comparison of leukocyte destruction between patients with acute onset (red) and non-acute onset (blue). *, P < 0.05. NS, not significant (P > 0.05).

For etiological associations, the mean proportion of leukocyte destruction in Phase 1 was higher in patients with non-infectious acute illness (7.9 ± 3.2 %, n = 4) compared to those with infectious etiology (4.0 ± 0.6 %, n = 26), though this difference did not reach statistical significance (p=0.07, Fig 2B).

For onsets, patients with acute onset exhibited significantly higher leukocyte destruction (7.0 ± 1.8 %, n = 7) compared to those with non-acute presentation (3.7 ± 0.6 %, n = 23, p < 0.05, Fig 2C). Among sepsis cases, a similar trend was observed but did not reach statistical significance.

### Additional Morphological Features in Acute and Non-acute Populations

To investigate whether other leukocyte parameters at Phase I differed between the onset-differentiated groups, a detailed analysis was conducted (Table 1). Promyelocytes, myelocytes, and metamyelocytes were observed at higher levels in the non-acute group; however, this difference was not statistically significant. In contrast, for toxic granulation, the proportion of patients demonstrating this feature was significantly higher in the non-acute group at 70%, compared to 14% in the acute group (p<0.01).

**Table 1.**
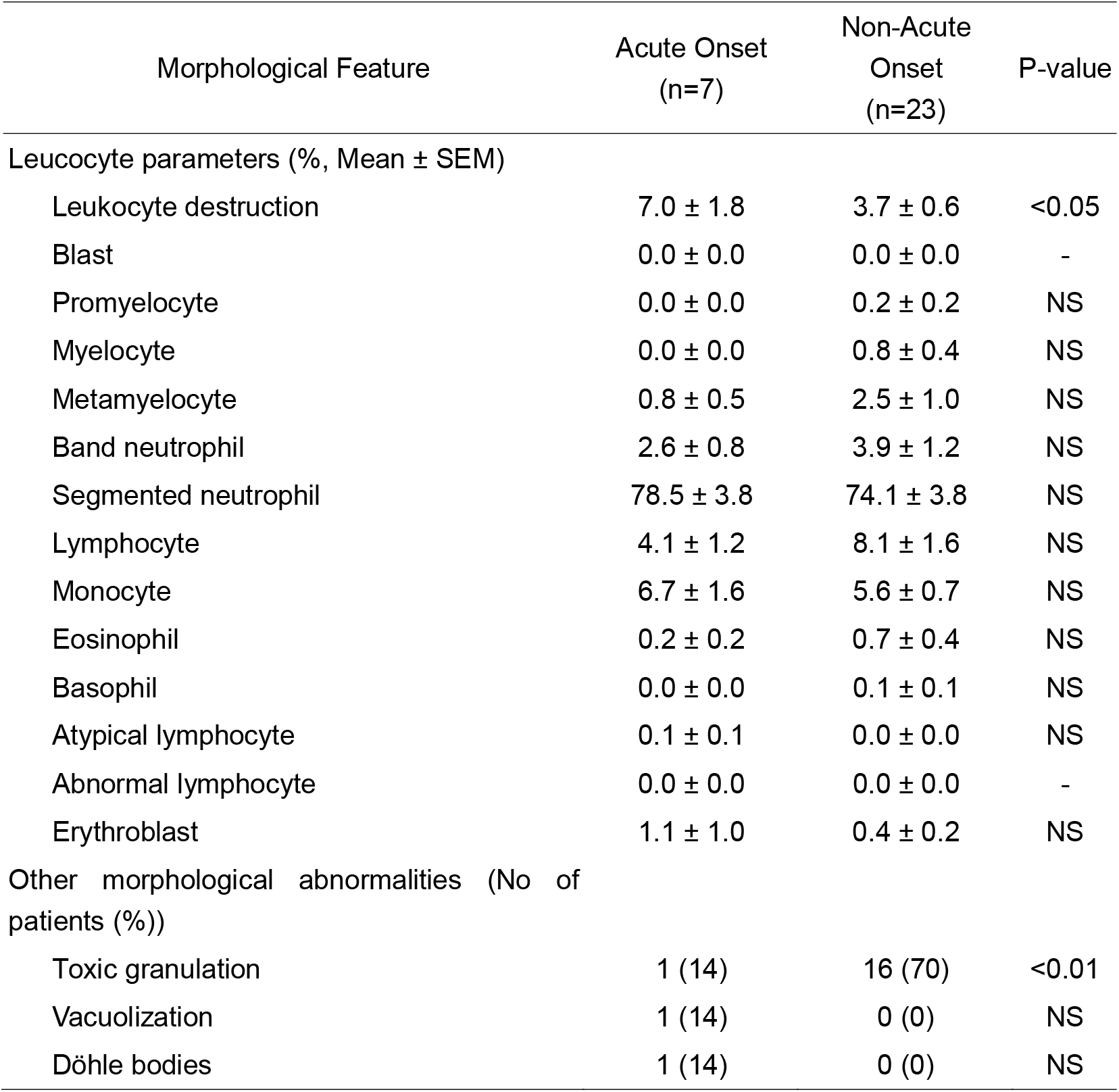
Leukocyte morphological features in patients with acute versus non-acute onset.

| Morphological Feature | Acute Onset<br>(n=7) | Non-Acute<br>Onset<br>(n=23) | P-value |
| --- | --- | --- | --- |
| Leucocyte parameters (% Mean ± SEM) | | | |
| Leukocyte destruction | 7.0 ± 1.8 | 3.7 ± 0.6 | <0.05 |
| Blast | 0.0 ± 0.0 | 0.0 ± 0.0 | - |
| Promyelocyte | 0.0 ± 0.0 | 0.2 ± 0.2 | NS |
| Myelocyte | 0.0 ± 0.0 | 0.8 ± 0.4 | NS |
| Metamyelocyte | 0.8 ± 0.5 | 2.5 ± 1.0 | NS |
| Band neutrophil | 2.6 ± 0.8 | 3.9 ± 1.2 | NS |
| Segmented neutrophil | 78.5 ± 3.8 | 74.1 ± 3.8 | NS |
| Lymphocyte | 4.1 ± 1.2 | 8.1 ± 1.6 | NS |
| Monocyte | 6.7 ± 1.6 | 5.6 ± 0.7 | NS |
| Eosinophil | 0.2 ± 0.2 | 0.7 ± 0.4 | NS |
| Basophil | 0.0 ± 0.0 | 0.1 ± 0.1 | NS |
| Atypical lymphocyte | 0.1 ± 0.1 | 0.0 ± 0.0 | NS |
| Abnormal lymphocyte | 0.0 ± 0.0 | 0.0 ± 0.0 | - |
| Erythroblast | 1.1 ± 1.0 | 0.4 ± 0.2 | NS |
| Other morphological abnormalities (No of patients (%)) |  |  |  |
| Toxic granulation | 1 (14) | 16 (70) | <0.01 |
| Vacuolization | 1 (14) | 0 (0) | NS |
| Döhle bodies | 1 (14) | 0 (0) | NS |

## Discussion

The present study is likely the first to systematically quantify leukocyte destruction morphology in the peripheral blood of patients with suspected sepsis and acute systemic illness, and to correlate these findings with clinical course and outcome. While prior research has established the importance of NETs and neutrophil activation in the pathogenesis [1, 6-8], no studies were found that explored the practical utility of routine blood smear analysis for detecting these phenomena.

Morphological destruction of leukocytes is sometimes observed in peripheral blood smears and has been conventionally recognized as an artifact related to sample storage conditions or smear preparation [9-10]. A report suggests that it is time-dependent, rapidly increasing after 6 hours [10]. However, this duration is considerably longer than the typical time taken for routine smear preparation in the present study. Furthermore, while that report evaluates what they term “Smudge cells,” the images presented do not resemble the leukocyte destruction defined in our study, suggesting they might be observing different leukocyte morphologies. While mechanical forces during smear preparation could potentially induce leukocyte destruction, it is evident that, at least with acute systemic illness, many cells are vulnerable to such changes.

Our findings indicate that the morphology of leukocyte destruction in acute systemic illness, particularly during the early stages of critical illness, can reflect the patient’s clinical background. The lack of association with short-term mortality suggests that leukocyte destruction is not a direct prognostic marker. However, the findings regarding leukocyte destruction and onset suggest a potential utility as a diagnostic tool, and if this leads to the development of appropriate interventions, it might also influence patients.

The observation of leukocyte destruction in cases of severe illness aligns with recent reports showing that morphologically similar NETs can be induced by a wide range of stimuli [3, 5, 6, 11]. Since the excessive NETs formation has been implicated in microvascular thrombosis, organ dysfunction, and poor outcomes in sepsis and COVID-19 [4, 5, 12, 13], leukocyte destruction in routine clinical laboratory practice should also be thoroughly investigated for similar pathological conditions and outcomes.

Another significant finding is the prevalence of leukocyte destruction morphology in cases with acute onset, alongside a higher proportion of toxic granulation in patients with non-acute onset. While the appearance of toxic granulation is classically known to occur during severe inflammation [14] our study reveals a novel insight: the time from disease onset also plays a role in its manifestation. Although changes in the proportion of toxic granulation over time have been observed previously [15], its potential dependence on the onset timing was not previously recognized. Furthermore, while not statistically significant, the observed lower proportion of leukocyte destruction in infectious cases compared to non-infectious acute illness might be attributed to infectious patients often presenting to the ICU at a more advanced stage of disease progression. The discovery of a time lag between leukocyte destruction and the appearance of toxic granulation suggests the presence of distinct leukocyte populations with varying potentials to elicit these responses. This provides a crucial insight: there might be unidentified leukocyte maturation stages, or novel differentiation pathways, yet to be fully understood.

Our study has several limitations, including the small sample size, retrospective design, and reliance on manual morphological assessment, which may introduce observer variability. Future research should validate these findings in larger, prospective cohorts and incorporate advanced imaging and molecular techniques to better characterize the mechanisms and clinical implications of leukocyte destruction and NETs formation.

## Conclusion

Leukocyte destruction in acute-onset cases and toxic granulation in non-acute-onset cases show promise as diagnostic indicators. Further research is needed to clarify their clinical significance and biomarker potential in critical care.

## Data Availability

All data produced in the present work are contained in the manuscript

## Abbreviations

ICU: intensive care unit
NETs: neutrophil extracellular traps
COVID-19: Coronavirus disease 2019
EDTA-2K: ethylenediaminetetraacetic acid dipotassium salt dihydrate

## Acknowledgement

We extend our sincere thanks to Haruka Obayashi, Minami Okamoto, and Yukimi Katagami for their invaluable assistance with sample preparation.

## Declaration of Generative AI and AI-assisted technologies in the writing process

During the preparation of this work, the authors used Perplexity (Perplexity AI) and Gemini (Google AI) in order to assist with sentence structuring and rephrasing for improved flow. After using these tools/services, the authors reviewed and edited the content as needed and take full responsibility for the content of the publication.

Leukocyte morphological features in phase 1 are shown. Statistical comparison for toxic granulation was performed using the Chi-squared test. SEM, standard error of the mean. NS, not significant.

